# A Comparative Analysis of Dementia Strategies of Fifteen European Countries in the Context of Glasgow Declaration and WHO’s Global Action Plan

**DOI:** 10.1101/2025.02.08.25321936

**Authors:** Smruti Bulsari, Nureen Izyani Hashim, Kiran Pandya, Russell Kabir

**Affiliations:** Institute of Public Health and Wellbeing, University of Essex, Colchester (Essex): UNITED KINGDOM; NIHR Applied Research Collaboration (ARC), East of England, Cambridge: UNITED KINGDOM; School of Allied Health, Faculty of Health, Medicine and Social Care, Anglia Ruskin University, Norwich: UNITED KINGDOM; Srimad Rajchandra Institute of Management and Computer Applications, Uka Tarsadia University, Bardoli (Gujarat): INDIA

## Abstract

Dementia prevalence across the globe is in alarming proportion and it is even expected to rise in the future. The World Health Organisation (WHO) had declared dementia as a health priority, way back in 2009 and had recommended then, that at least high-income countries develop a dementia action plan, and other countries develop a national dementia strategy (NDS). Later in 2014, European Countries came together to sign the Glasgow Declaration and agreed to develop their respective NDSs. Yet, a few countries still do not have their NDS. Moreover, some countries do not have their NDS in English. This study attempts to compare the dementia strategies of 15 European countries, which has a comprehensive NDS in English language. The study further examines how well these NDSs comply with the Glasgow Declaration and the WHO’s Global Action Plan guidance. We undertake cluster analysis to classify NDSs of these countries in terms of similarity in the content. We make use of word clouds to get an overall idea about the clusterwise contents of the NDSs, and then use algorithmic approach to content analysis for identifying the clusterwise key focus areas of these dementia strategies. We make a comparative analysis of these NDSs in the perspective of dementia prevalence, demographic profile, per capita gross domestic product (GDP) and predominant healthcare financing model. We have found that irrespective of the prevalence, country’s demographic profile, GDP per capita or the predominant financing model, dementia strategies primarily focus on “care”. We discuss the cost-effectiveness of prevention and person-centered care (PCC) and suggest according priorities to these in the future NDSs, as these are also the focus areas of Glasgow Declaration and the GAP.

## 1. Introduction

Dementia is a neurodegenerative disorder and a life-changing health condition that influences a person’s memory, cognition, language, and behaviour. It is progressive in nature and one of the major causes of disability among people aged 60 and above. Dementia affects not only the individual but everyone in the family, and more specifically, the main caregiver. Globally, it is the seventh leading cause of death (1). Dementia prevalence across the globe was 55 million in 2019, which is an increase from over 36 million in 2010, and is expected to rise to 135 million in 2050 (2,3). The World Health Organisation (WHO) declared dementia as a world health priority way back in 2009. WHO had also recommended high-income countries to develop a national dementia action plan and low and medium-income countries should develop national dementia strategies with a focus on improving primary healthcare and community services (4). Despite this, very few countries have a national action plan or a national dementia strategy (NDS). WHO later, launched a Global Action Plan (GAP) with an objective to improve the lives of people living with dementia (PLwD) and their caregivers, as well as reduce the impact of dementia by focusing on preventative measures (5).

European Countries came together to sign the Glasgow Declaration in 2014 and agreed to develop their respective NDSs. Though, France, Cyprus, Denmark, Belgium (Flanders as well as Wallonia), Israel, Norway and Scotland had dementia strategies even before 2014. It may be noted that while Israel is geographically in Asia, it is an associated state of the European Union and is also a member of Alzheimer’s Europe. Also, some of these countries revised their existing NDSs after the Glasgow declaration.

Glasgow Declaration was launched at the 24th Annual Conference of the Alzheimer Europe, (2014), and it was unanimously adopted by 26 member countries. Alzheimer Europe is a non-profit, non-governmental organisation (NGO) comprising 41 national Alzheimer Association and 36 countries of Europe. It works towards improving lives of people living with dementia and their caregivers, through change in policy and practice, as well as changing society’s perceptions to combat stigmatising dementia. However, 17 out of 41 European countries still do not have an NDSs. England refers to it as Prime Minister’s Challenge to Dementia, Finland as the Memory Programme, and Belgium, Cyprus, Greece, Norway, and Wales as Action Plans. NDSs are available in the public domain on the Alzheimer Europe’s website (6), from where we have downloaded for this study. We will refer to all national-level dementia strategies / action plans, by whatever name they may be referred to, as “National Dementia Strategy(ies)” or “NDS”.

There is a dearth of scientific literature analysing the contents and coverage of these dementia strategies. Some literature on comparative analysis of dementia strategies could be found in the context of Canada and its provinces (7), or in general, countries across the globe (8), also in the context of the future NDS of Canada. Dementia Strategies of 29 countries, published on the website of Alzheimer’s Disease International (9).

Other comparative analyses literature focus on specific areas like palliative care (10), person-centered care (PCC) (11), overall care (12), interaction between public health systems and social care systems, coordination in transition from home to residential care settings, and dementia care professionals (13) or human rights (14). Alzheimer Europe already has a system of doing comparative analysis of dementia strategies their member countries (15,16).

This study attempts to contribute to the scientific literature by undertaking a comparative analysis of the contents of NDSs of European countries and examine the extent to which these comply with the fundamental rights laid down in the Glasgow Declaration, as well as the key action areas of the WHO’s Global Action Plan. However, we have included only those countries which have their NDS in English.

This paper is organised into five sections, with sub-sections: Sub-section 1.1 describes the fundamental rights laid down in the Glasgow Declaration and the key action areas of the GAP of the WHO. This builds a perspective within which, this study compares dementia strategies of selected European countries. Section 2 provides the rationale for selection of the Alzheimer’s Europe member countries for comparative analysis. Sub-section 2.1 discusses the framework used in this study, for undertaking a comparative analysis of dementia strategies, and explains the Natural Language Processing (NLP) approach to comparative analysis. Section 3 describes the method of classification of NDSs into different categories and presents some basic characteristics of the country in terms of population, health and income. Clusterwise major highlights of the dementia strategies is described in Sub-section 3.1. Sub-section 3.2 highlights the key themes for each cluster, using content analysis. Section 4 discusses the findings and provides recommendations for the future NDSs, in the context of Glasgow Declaration and GAP, along with their cost considerations. Section 5 concludes.

### 1.1 Fundamental Rights of Glasgow Declaration and Key Action Areas of the GAP

Glasgow declaration called upon devising an umbrella dementia strategy of the entire Europe and also individual national dementia strategies of the member countries (17).

The fundamental rights laid down in the Glasgow Declaration are, the rights to:

- timely diagnosis,
- access to post-diagnostic support,
- person-centric, and coordinated care,
- equitable access to treatment and therapeutic interventions, and
- respect as an individual in their community.

GAP has listed seven key action areas:

- Dementia as a public health priority,
- Developing awareness about dementia and taking dementia-friendly initiatives,
- Reduce the risk of dementia incidence,
- Timely diagnosis, support, treatment, and care,
- Providing necessary support and training to the caregivers of people with dementia,
- Develop a core set of dementia indicators and collect data on those indicators, every two years, and
- Undertaking research and focusing on innovations to improve the lives of people living with dementia.

The GAP is expected to provide a blueprint for governments to improve the quality of life of PLwD and their caregivers. Although the GAP is not legally binding, the ministries of all member states are obliged to provide regular progress reports (three times over seven years) to the WHO (18). Arthurton et. al. (19) observe that regular monitoring of the work has resulted in more countries coming up with dementia strategies.

There is a dearth of literature examining the extent to which, the NDSs are informed by the Glasgow Declaration or the GAP. This study aims to bridge that gap by providing evidence on the extent to which the NDSs of these 15 European countries address the rights laid down in the Glasgow Declaration and the key action areas of the GAP.

## 2. Methods: Selection of the Countries, Framework and Approach

Alzheimer Europe website (6) lists 41countries and 45 clickable links. England, Scotland, Wales and Northern Ireland are counted as one, though there are separate dementia strategies for each of these countries. Similarly, Belgium (Flanders and Wallonia) is counted as one, though they have separate dementia strategies for each region. As discussed, 17 European countries (Bosnia and Herzegovina, Bulgaria, Croatia, Estonia, Hungary, Jersey, Latvia, Lithuania, Montenegro, North Macedonia, Poland, Romania, Serbia, Slovakia, Turkey and Ukraine) do not have a dementia strategy, whereas Armenia and Portugal do not have a comprehensive dementia strategy that can be included for comparative analysis. Out of 22 countries, 11 countries (Belgium, Czech Republic, France, Iceland, Italy, Luxembourg, Spain, Sweden and Switzerland) have NDSs in languages other than English. United Kingdom has separate NDS for England, Scotland, Wales and Northern Ireland. Thus, a total of 15 NDSs are analysed. It would be important to state that out of the 15 NDSs selected for our study. England, Belgium (Flanders), Gibraltar, Norway and Scotland have revised their NDSs over time. We have included only the latest dementia strategies for our analysis. This is because, we are interested in making a comparison of the ongoing steps undertaken by different countries, for PLwD, and for the prevention of dementia.

These 15 countries, whose NDSs are included in this study are: Austria, Belgium, Cyprus, England, Finland, Germany, Gibraltar, Ireland, Israel, Malta, Netherlands, Northern Ireland, Norway, Scotland and Wales.

### 2.1 Framework and Approach: Comparative Policy Analysis

Comparative policy analysis is concerned with examining and understanding variations in the outcomes of governmental activities over time and jurisdictions. It attempts to answer questions related to the influence of social, economic and political situations on the contents and provisions of the public policies (20). This implies that if there are differences in socioeconomic and political situations across countries, it would be reflected in the differences in their respective public policies. Out of the 15 European countries in this study, 14 are members of the European Union (EU). Since EU directives promulgate policies in their member countries, we expect homogeneity across these NDSs. These countries are also member states of Alzheimer’s Europe. Therefore, we expect the NDSs of these countries to be guided by the fundamental rights laid down in the Glasgow Declaration of Alzheimer Europe (17).

We employ the *comparative method* in this study, which helps designing a framework for describing the content, timings and outcomes of policies of different countries (21). Peters (22) suggests the use of this method for *prima facie* classification of similar policies in groups. We explore similarities and differences in the policy text using natural language processing (NLP) tools. There is a growing acceptance and use of NLP tools for policy analysis (23–25).

We apply k-means cluster analysis to classify the NDSs based on similarities (and dissimilarities) in their contents. Lie et al. (26) suggest the use of k-means cluster over other methods of classification for its efficiency.

We then generate word frequencies to examine the focus areas of dementia strategies in each of these clusters. Word frequencies are often represented as word clouds, which is one of the static methods of summarising the text (27); larger font size represents higher frequencies. While single words would give a broad idea about the focus of dementia strategies, using a set of two (or three) consecutive words, called bigrams (or trigrams) in NLP terminology, would help contextualise the use of words (28). Details of pre-processing the text for required contextualisation is given in supporting information (Appendix – A). This process would be similar to undertaking content analysis, except that we use the algorithmic technique, coded in Python 3.11. These bigrams and trigrams would then be used to identify themes, by grouping words with similar meanings used in similar context, to get a contextual idea of the themes in the dementia strategies’ clusters.

The advantage of using NLP techniques is that a large quantum of text can be analysed without the possibility of missing or overlooking any part of the text. It eliminates biases that might enter while interpreting the text manually. Moreover, manual interpretation of texts usually ranges over days, sometimes even with some breaks, which might lead to a change in the context of interpreting the text, leading to comparability issues. There is no subjectivity involved when using the NLP tools, and the algorithm takes several minutes to several hours, depending upon the quantum of text and the processor specifications. The disadvantage on the other hand is, since these techniques are based on word frequencies, the words with very low frequencies would not appear, despite being there in the text.

Different European countries have different dementia prevalence as well as different models of financing of healthcare services. Most countries adopt the Beveridge model, while the others follow either the Bismarck model or a mix of the two. Healthcare is financed by the government through tax payments in the Beveridge model, whereas it is financed by the insurance system, with joint contribution of the employers and the employees, in Bismarck model (29). This study examines the patterns in these characteristics of the countries, if any: dementia prevalence, population, percentage of population above 65 years of age, life expectancy at birth, per capita income and their healthcare financing model, to understand whether these characteristics determine the contents of their respective dementia strategies.

## 3. Results: Dementia Strategies Classification

The data for this study is the corpus of dementia strategies of selected countries of the European Union. Dementia strategies are downloaded from Alzheimer Europe (6), using a web-scraping algorithm developed by SB in Python 3.11.

Dementia strategies across these European countries are cleaned and sentence tokenised before applying the k-means cluster analysis algorithm. Cleaning refers to removal of header / footer text, table of contents, tables containing numbers and other text that can bias the analysis. The process of separating each sentence of the text corpus is called sentence tokenising. We then subject these sentences to the algorithm, Elbow method, to identify the optimal number of clusters “k” in which these NDSs will be categorised. Elbow is one of the most commonly used methods of determining the optimal clusters and provides a visual representation (30,31). The output of the Elbow method in Fig 1, suggests four clusters. Thus, k is specified to be equal to 4 to classify the NDSs of these 15 countries.

**Fig 1:**
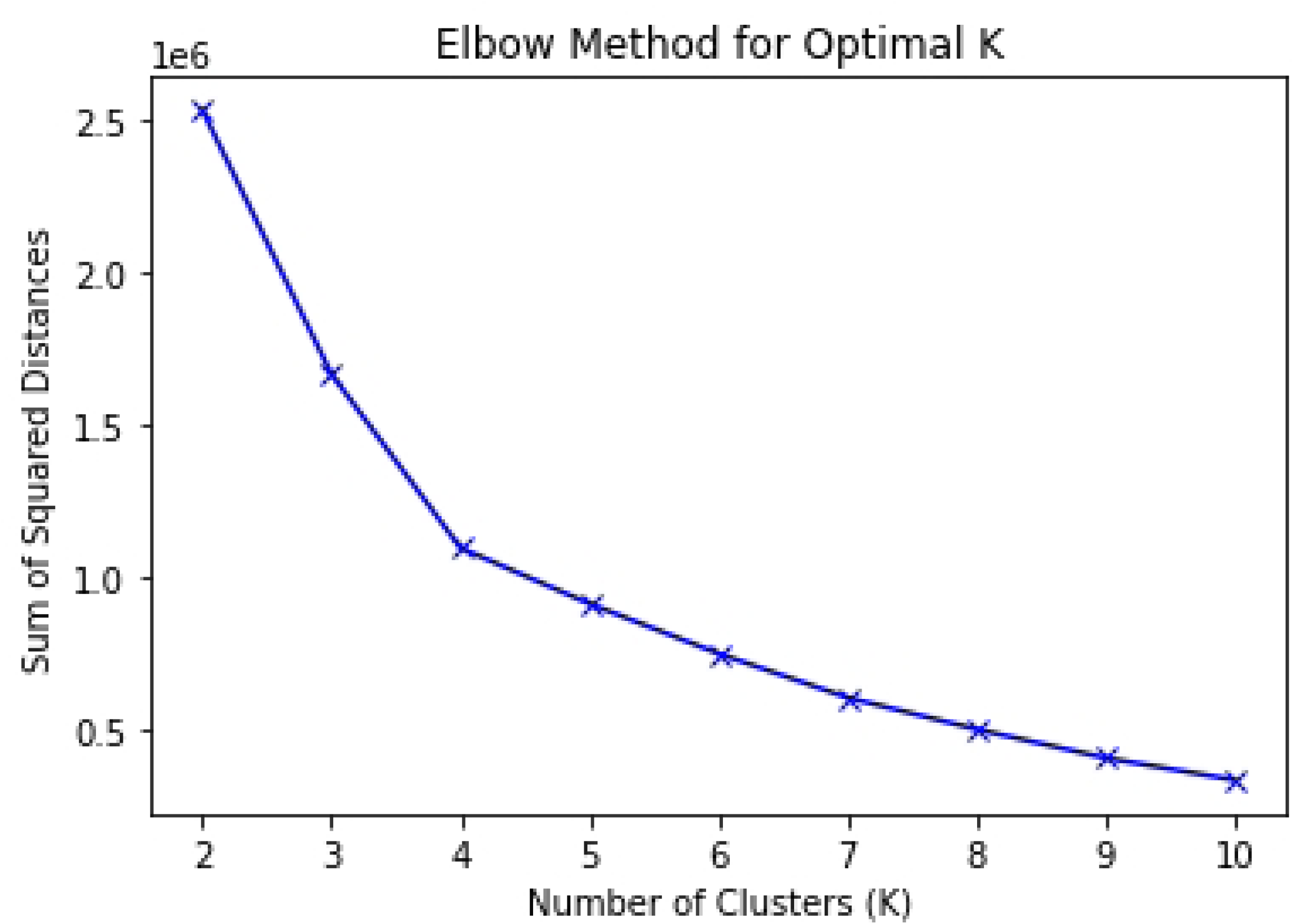
Sum of Squared Distances for Different Values of “k”: Elbow Method The elbow appears at k=4, indicating the optimal number of cluster to be four.

These four clusters, with the countries’ cluster membership, are shown in Table 1, where, the countries within each cluster have similarities in the contents of their NDSs:

**Table 1:** Cluster-Membership of Countries based on their NDSs contents.

| Cluster No. | Countries | Number of Countries in each cluster |
| --- | --- | --- |
| 1 | Austria, Netherlands, Israel, Scotland, Gibraltar, Wales, Denmark, Cyprus, Belgium and Finland | 10 |
| 2 | Germany | 1 |
| 3 | Norway | 1 |
| 4 | England, Ireland, Malta, Northern Ireland and Greece | 5 |

Table 1 shows that cluster 2 and 3 are single country clusters comprising Germany and Norway respectively. We then examine the country characteristics to understand the reasons for similarities and differences in the dementia strategies. We examine patterns in population, percentage of population above 65 years of age, life expectancy at birth, per capita income, and dementia prevalence across these clusters. The summary measures of these indicators are shown in table 2, and the details on these indicators for each of these 15 countries are given in Table s1 (supporting information, Appendix – B). It may be noted that the data on these indicators are available as consolidated figures for all the constituent countries of the United Kingdom. It was possible to find the data, separately for these countries, but there were comparability issues and hence, we decided to include consolidated figures only. We have excluded these figures in estimating the summary measures, because Scotland and Wales are in cluster 1, whereas England and Northern Ireland are in cluster 4.

**Table 2:**
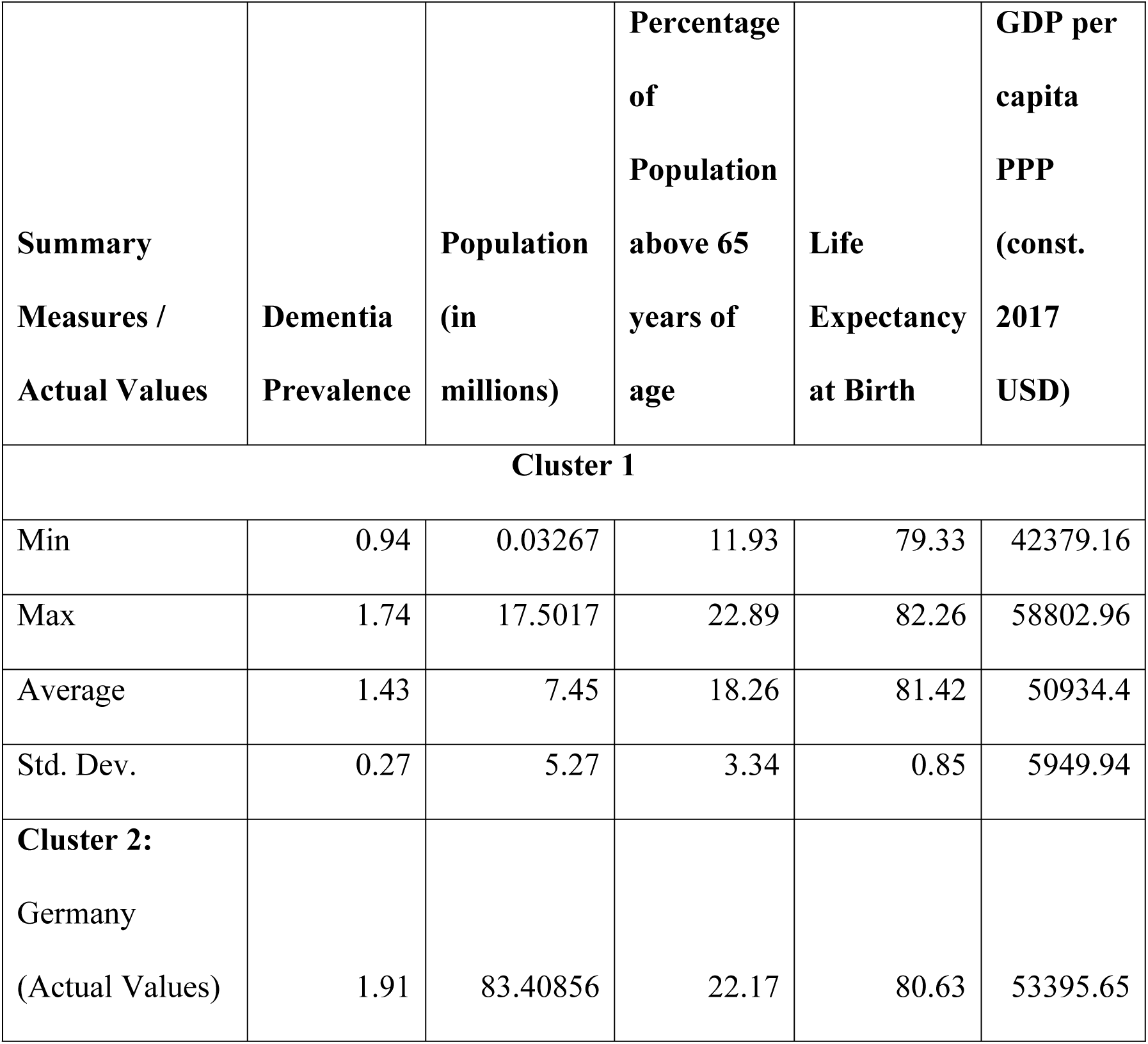
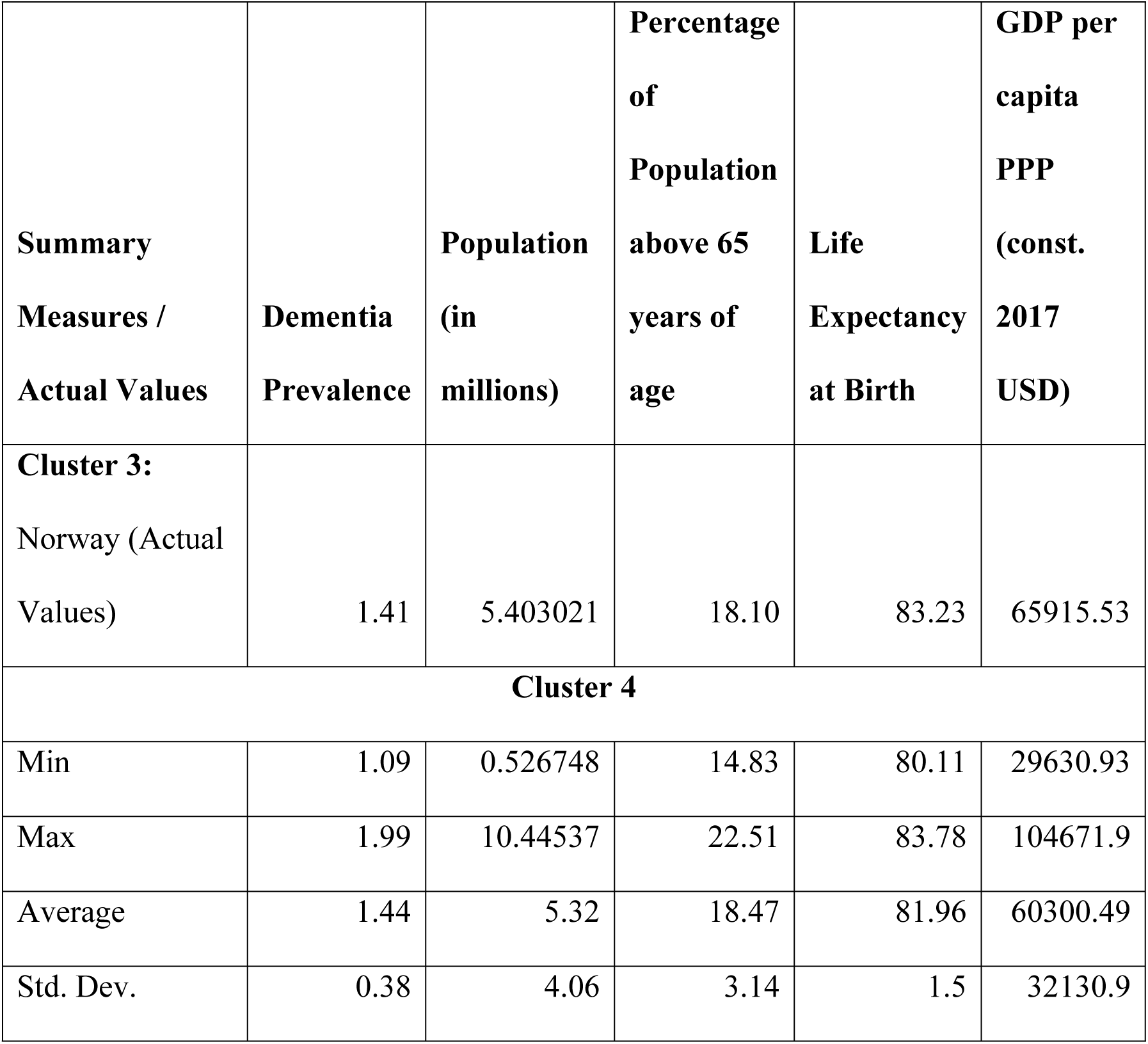
Clusters and Summary Measures of Dementia Prevalence, Demographic Profile and GDP Per Capita.

Table 2 shows that Germany, the only country in cluster 2, differs from the rest of the clusters in terms of having a very high dementia prevalence and having the highest population among all the 15 countries included in this study.

Norway, the only country in cluster 3 differs from the rest of the clusters in terms of highest per capita GDP, among the countries included in this study.

There is no apparent difference in the country profile indicators for clusters 1 and 4. Therefore, we examined the patterns in healthcare financing system in each of these countries (supporting information, Table s1, Appendix – B).

Eight out of 17 countries have adopted the Beveridge model of healthcare financing, and five have adopted the Bismarck model. The Netherlands follows a mix of Bismarck and private voluntary insurance. Cyprus follows a hybrid of Bismarck and Beveridge models; it is a mix of government funding and mandatory social insurance contributions (32). The Gibraltar Health Authority uses a healthcare model closely linked with that of the National Health System (NHS) in the United Kingdom, thereby following the Beveridge model (33), and Israel follows the Bismarck model of healthcare financing (34).

Taking this information into account, 4 out of 10 countries of cluster 1, follow the Bismarck model, 2 follow Bismarck + Beveridge (hybrid) and 5 follow the Beveridge model of healthcare financing. In cluster 4, 4 out of 5 countries follow the Beveridge model of financing; only Greece follows the Bismarck model. Thus, cluster 4 is largely homogeneous in terms of countries following the Beveridge model. This distinguishes cluster 4 from cluster 1.

### 3.1 Major Highlights of Dementia Strategies by Clusters

The highlights of dementia strategies of each cluster are represented in the form of word clouds in Fig 2 (a) to 2(d).

**Fig 2.**
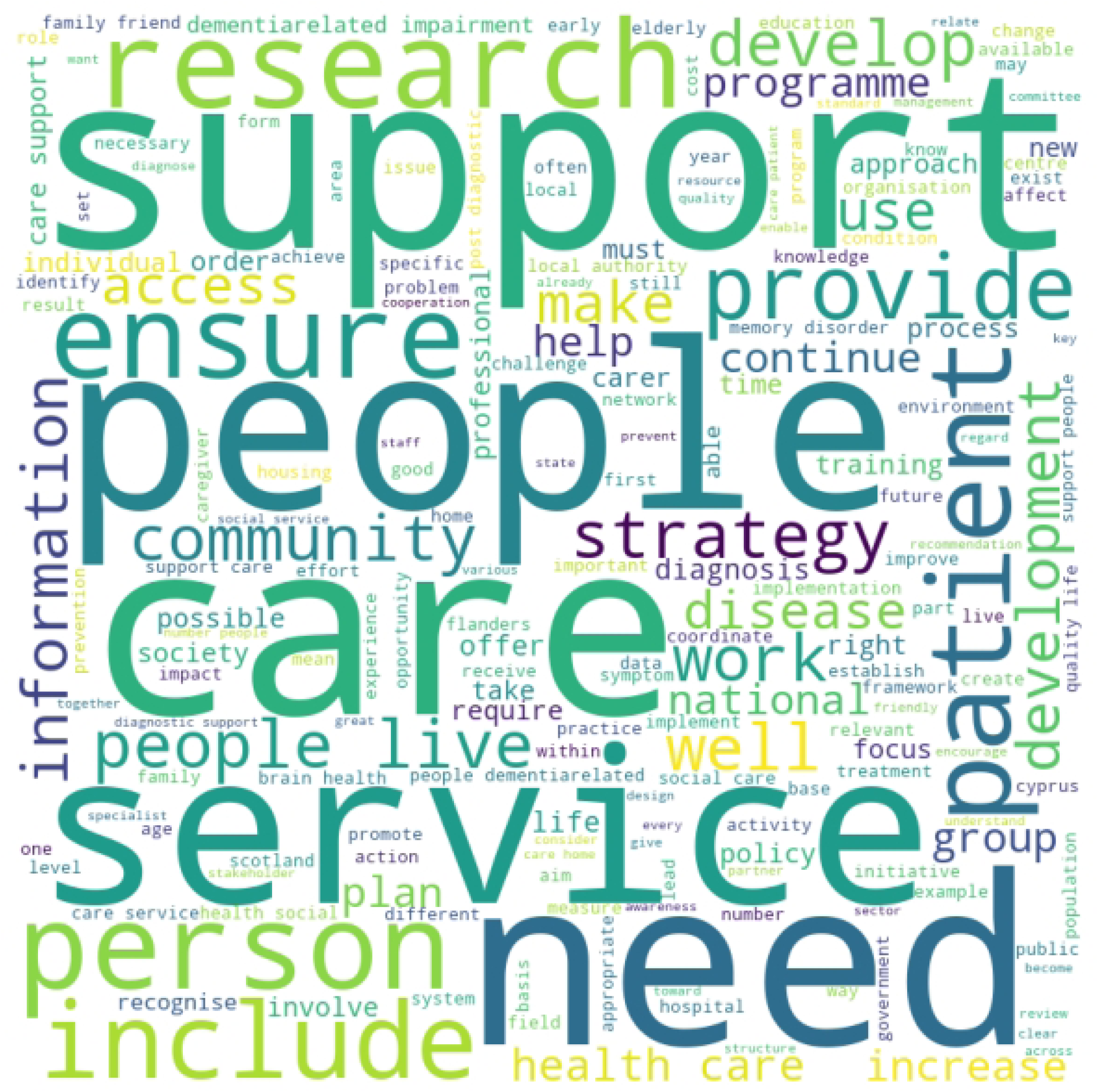
(a): Word Cloud for Dementia Strategy Text of Cluster 1 Countries Highlights these words in the combined text of dementia strategies of countries of cluster 1: “support”, “care”, “service”, “need” and “people”.

**Fig 2.**
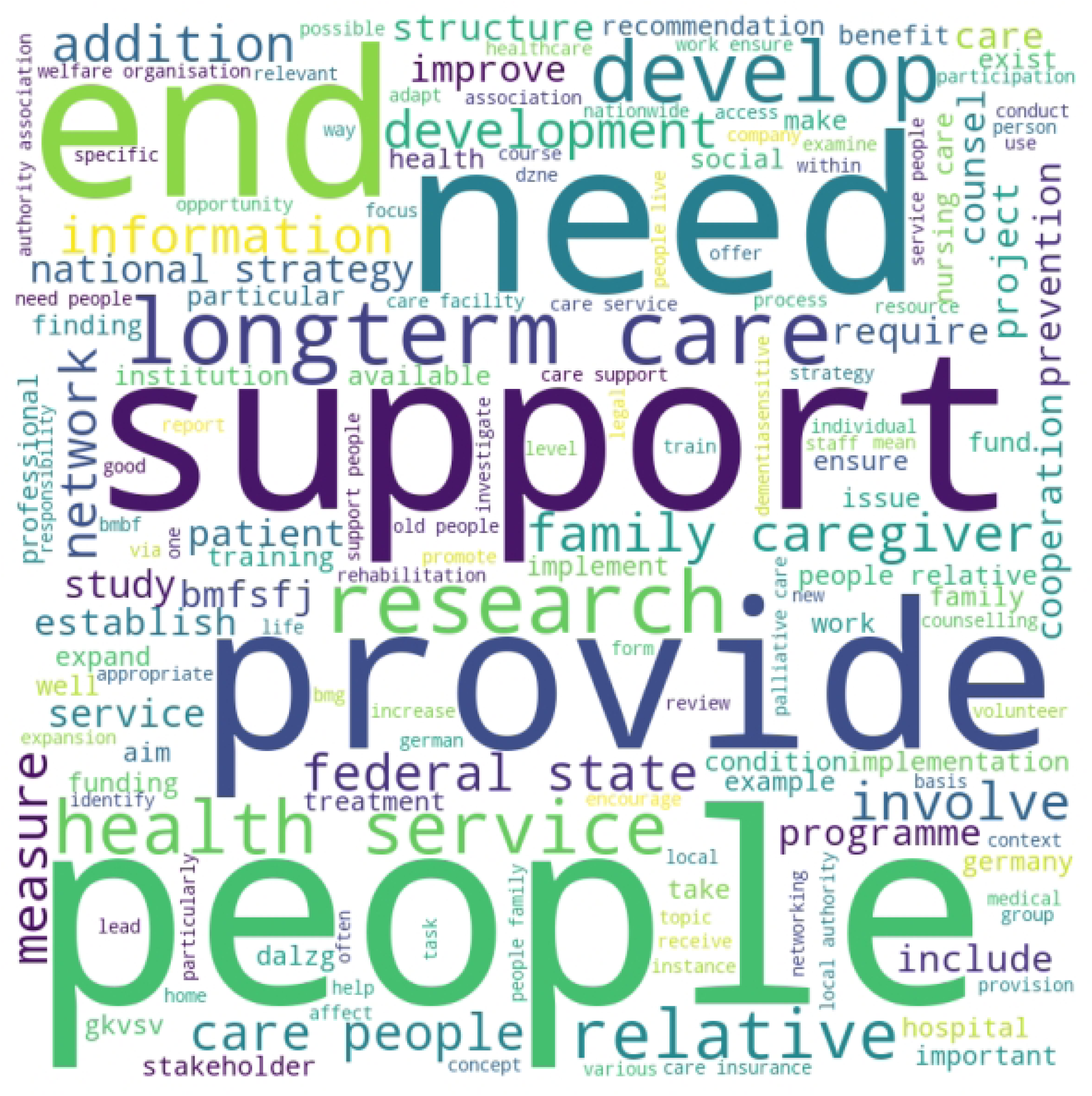
(b): Word Cloud for Dementia Strategy Text of Cluster 2 Country (Germany) Highlights these words in the national dementia strategy of Germany, the only country in cluster 2: “end”, “need”, “support”, “provide” and “people”.

**Fig 2.**
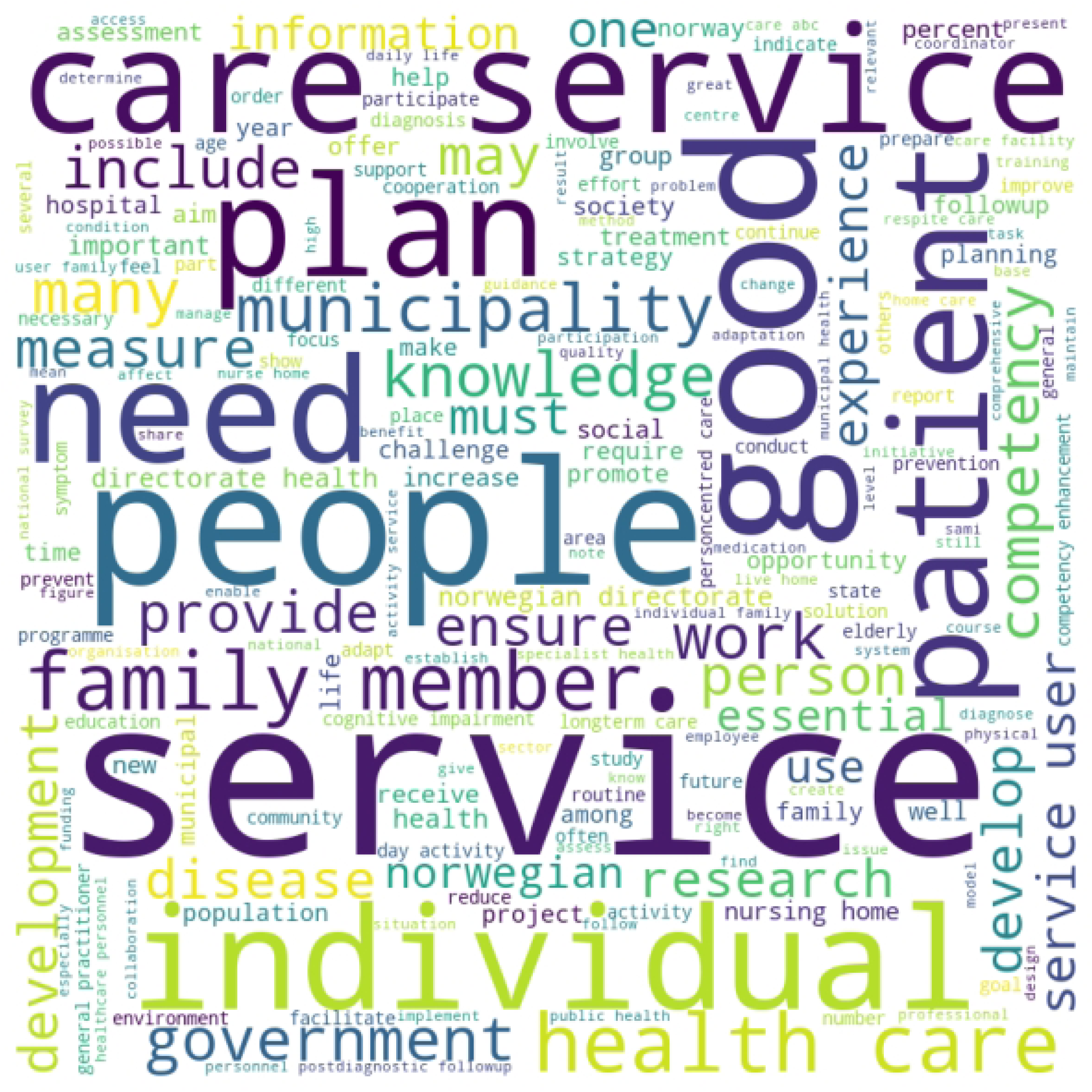
(c): Word Cloud for Dementia Strategy Text of Cluster 3 Country (Norway) Highlights these words in the national dementia strategy of Norway, the only country in cluster 3: “care”, “service”, “people”, “need”, “good” and “patient”.

**Fig 2.**
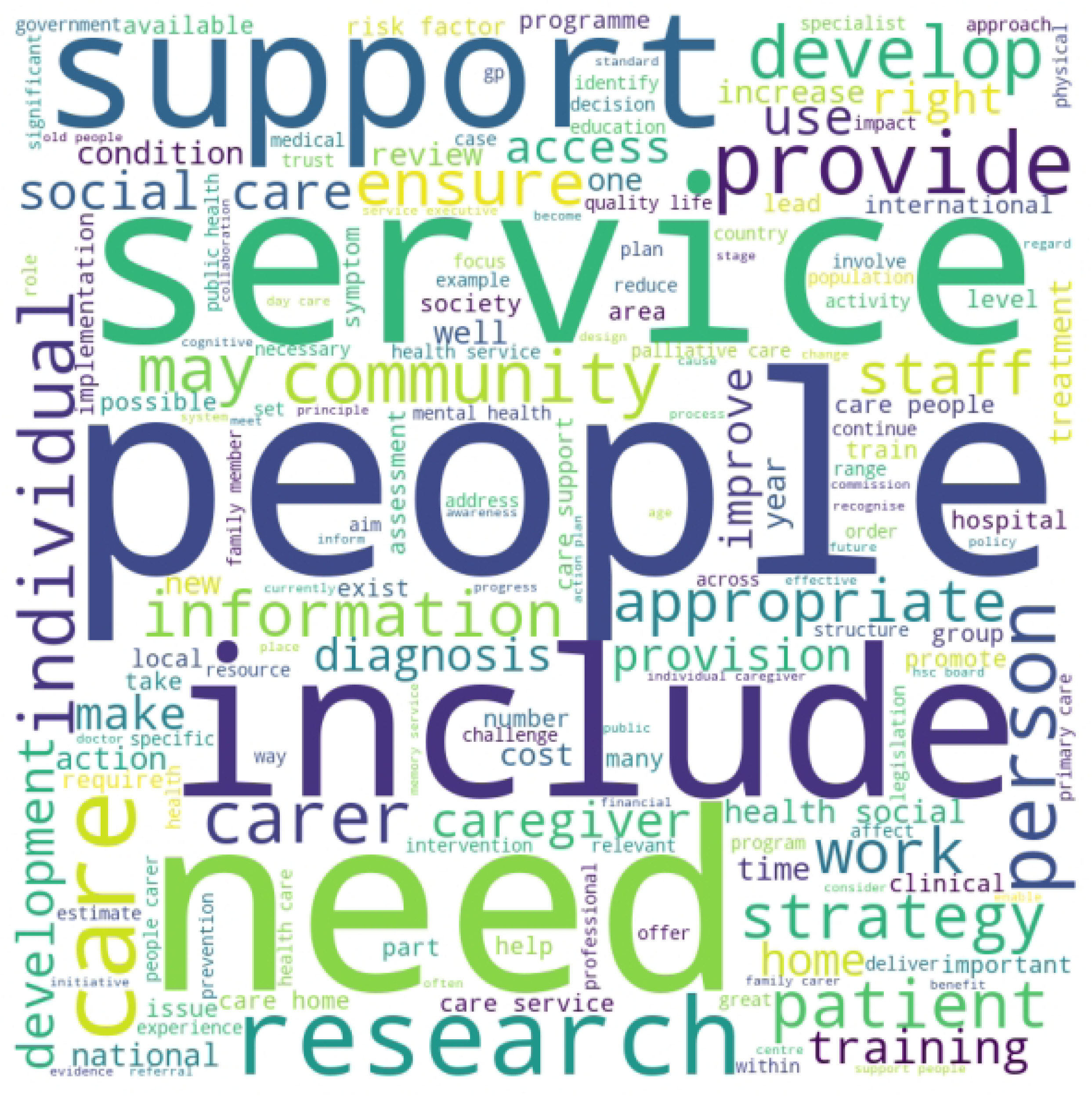
(d): Word Cloud for Dementia Strategy Text of Cluster 4 Countries Highlights these words in the combined text of dementia strategies of countries of cluster 4: “support”, “service”, “people”, “include” and “need”. The focus on “research” can also be seen in this word cloud.

The word cloud is based on the frequencies of the words occurring in the corpus of NDSs of each cluster. Word-level preprocessing of text is undertaken at this juncture for developing the word cloud and for further analysis. Preprocessing steps are explained in the supporting information (Appendix – A). Larger fonts, and repetition of words indicate higher frequency.

Fig 2 (a) shows that “support”, “care”, “service”, “need” and “people” are the top five frequently occurring terms in the combined text of the dementia strategies of cluster 1. Terms “support” and “service” can be interpreted to a part of the broader concept “care”. These five terms together would mean the focus of these dementia strategies is on “care for people (with dementia) in need”. Since “dementia” and “Alzheimer’s Disease” are removed from the list of words to be counted, they do not appear in the word cloud. The context is nevertheless, “people with dementia” and hence it is mentioned in the brackets.

Fig 2 (b) highlights the terms “end”, “need”, “support”, “provide” and “people”. The term “support” is interpreted as synonymous to “care”, and the term “end” appears in the context of end-of-life. Therefore, the focus of the dementia strategy of Germany is on “providing end-of-life support to people (with dementia) in need”.

Norway has a dementia action plan. Unlike strategies, action plans have targets set with deadlines. Therefore, the term “plan” is one of the frequently occurring terms Fig 2 (c). The top frequently occurring terms are “care”, “service”, “people”, “need”, “good” and “patient”. This could be interpreted as “planning care for people and patients (with dementia) in need”. The term “patient” is largely used to express illness or a person who needs consultation from a general practitioner or hospital services. On the other hand, dementia is a condition, though PLwD might require hospital care at advanced stages.

Fig 2 (d) also highlights similar terms as the other clusters: “support”, “service”, “people”, “include” and “need”. There is some emphasis on “research” too, in this word cloud. This can be interpreted as “care for people (with dementia) in need” and “(dementia) research”.

All these word clouds show that “care for people with dementia” is the core focus area of all NDSs, irrespective of the cluster in which they belong to. However, additionally, the dementia strategy of Germany emphasises on end-of-life care and the Norway Action Plan provides for care for patients, who are living with dementia. Cluster 4 countries emphasise on research, over and above “care for people with dementia”.

This implies that dementia strategies focus on post-diagnostic care, which is one of the fundamental rights of people living with dementia, as laid down in the Glasgow Declaration, and also one of the key action areas of the GAP.

This analysis is, however, based on individual terms. Contextualisation of these terms with the terms will give deeper insights into the key themes of dementia strategies.

### 3.2 Key Themes of Dementia Strategies Across Clusters

Content analysis is undertaken to identify clusterwise key themes of dementia strategies. We adopt conventional content analysis of identifying the frequency of the text, coding the text, and then identifying the key themes. The pre-processed text is used to generate bigrams and trigrams. Then frequencies of occurrence of these bigrams and trigrams in the dementia strategy text are determined and a list of the top 30 bigrams and 20 trigrams is prepared, in the descending order of their frequency.

Bigrams with similar meanings are combined into one theme; trigrams are combined in the same manner. For example, for cluster 1 countries, the bigrams containing the terms “care”, “support” and “service” are treated as synonymous for identifying the key themes. These terms are found together with “health”, “social”, “people”, “home” and “patient”. Therefore, this theme is labelled broadly as “care for people with dementia”. The frequency of occurrence of bigrams and trigrams in the clusterwise text of dementia strategies, and clubbing of terms therein, can be found in the supporting information (Tables s2 and s3, Appendix – C).

Bigrams’ frequencies reveal the following themes:

Cluster 1 (formed of 10 countries):

- Care for people with dementia, ranging from health care, social care, patient care, personal care and informal care. It may be noted that service is also interpreted as care in this context.
- People (with dementia), their family and friends
- Quality of life of people with dementia and related impairment

Cluster 2 (Germany):

- Care at different levels, for people with dementia: This includes long-term care, family care, nursing service, palliative care, short-term care, and care homes.
- Role of the State / system: This includes the role of local authority, system of (health-related) insurance and other organisations for the welfare of people living with dementia.
- Services for people living with dementia, which include health services, counselling and providing information.

Cluster 3 (Norway):

- Care at different levels, for people living with dementia: This includes long-term care, person-centred care, respite care, and care facility.
- Role of family members in care for people living with dementia.

Cluster 4 (formed of four countries):

- Care at different levels, for people living with dementia: social care, primary care, health care, daycare, family care, individual care, professional care and care management.
- Services related to public health.

Trigrams are interpreted in a similar manner as bigrams, except that these are chunks of three words. For an example, in cluster 1, trigrams containing the terms “care”, “support” and “service” are considered synonymous. They are found clubbed with the terms “health”, “social”, “people”, “live”, and “partners unpaid”. This theme too is therefore labelled as “care for people with dementia”.

One may notice the frequency of trigrams is less as compared to that of bigrams. This is because as the contextualisation becomes more specific, the probability of repetition of chunks of words reduces. However, bigger chunks of words (trigrams) provide more specific contextualisation, and therefore, it is important to analyse trigrams as well. Trigrams’ frequencies reveal the following themes:

Cluster 1:

- Care for people with dementia, especially social care and unpaid care, remains at the top of the themes of these dementia strategies.
- This is followed by dementia-related impairment, and its effect on family and friends.

Cluster 2 (Germany):

- There is a strong emphasis on long-term care for people with dementia.
- This is followed by services and research related to health, hospice, and palliative care facilities. There is also a focus on the need for defining the term “care”.

Cluster 3 (Norway):

- Care for people with dementia at different levels: This includes health care, specialist services, municipal health care, long-term care facility, home care, respite care, daycare, and services to patients.
- The focus is on the State’s system of healthcare. The Norwegian Dementia Action Plan focuses on having a national guideline, setting up a competency enhancement plan, and setting up a national advisory unit to facilitate services for people living with dementia.

Cluster 4:

This cluster has only one theme, which is care for people with dementia. This includes social care, family care, professional care, individual care, end-of-life care, daycare, and hospital care.

The results of bigrams and trigrams analysis show that “care” for people with dementia is the key theme across all clusters. The clusters differ from each other in terms of subsidiary themes like dementia-related impairment and, the role of the State in enhancing the access and services for people with dementia.

## 4. Discussion

Success of any strategy depends upon the cost of dealing with the illness / condition. It will be very instructive to understand different measurable and non-measurable costs of dementia. Understanding direct and indirect costs will also be important in this context. There are significant regional variations in the types of costs. For example, the United States of America (USA) has higher proportion of direct costs compared to that in European countries (35–37). We discuss the long-term cost effectiveness of prevention and person-centered care and recommend including the same in the future dementia strategies.

Focus on prevention would have long-term benefits on costs of care and burden on healthcare staff and professionals. Multidomain interventions like diabetes prevention, behavioural change, food supplements programme, reducing cardiovascular risk, to name a few, are found to reduce dementia risk; The Lancet Commission has identified 12 modifiable risk factors to prevent dementia(38), two additional factors are identified in 2024 (39). Braun et al. (40) undertake a systematic review of seven studies and summarise that the mean incremental quality of life-years (QALY) per person of multidomain interventions range between 0.04 and 0.06, and the cost-effectiveness ratio is 80,427.97 EUR for the best possible intervention. Albeit, they also found that a bad intervention may even cost as high as 104,189.82 EUR. Wimo et al. (41) estimate the long-term cost-effectiveness of the Finnish Geriatric Interventions Study to Prevent Cognitive Impairment and Disability (FINGER) programme. They found the mean incremental QALY to be 0.043 per person and the cost-effectiveness ratio between 17,000 Swedish Krona (SEK, equivalent to 2000 USD) and 600 SEK (equivalent to 70 USD) per person per year. McRae et al. (42) showed that people at risk of dementia, who were protected because of the changes made by them in their lifestyle resulted in a per person savings of 342 AUD; the average annual economic costs of dementia per person 35,550 AUD (including the indirect costs and the loss of employment for both, the PLwD and their caregivers). This makes a strong case for dementia strategies to focus on prevention.

GAP also instructs to give attention to person-centered care (PCC). Tay et al. (43) have estimate the incremental cost-effectiveness ratio of PCC for the Care for Acute Mentally Infirm Elders (CAMIE) unit for dementia in a hospital was lower than the threshold of 23,111 USD, and the incremental QALY was 0.045 per person. Little work is done to estimate the cost-effectiveness of PCC, though other outcomes like improvement in behavioural and neuropsychiatric symptoms, and care quality were observed for PLwD in acute care settings (44). Evans (45) suggests including the psychodynamic perspective for dementia diagnosis and post-diagnostic care, which provides an understanding into the resource constraints under which the healthcare professionals and care providers work, thereby providing for their mental health and wellbeing. This perspective in PCC could enhance the cost-effectiveness and QALY of PLwD. In the context of long-term care or hospital settings, Mohr et al. (46) have identified nine PCC interventions that could be useful for PLwD and their caregivers in organisational settings. These are: “real or simulated social contact, person-specific structured exercises, cognitive training, sensory enhancement, daily living assistance like providing the laundry services, help with activities of daily living (ADL), life-history oriented emotional support, training and support for professional caregivers, environmental adjustments (vision and sound), and care organisation”. Emphasis should also be on PCC in home care settings. Including the inputs of PLwD and their caregivers in devising dementia strategies would enhance the relevance and efficacy of NDSs’ provisions. Vinay & Biller-Andorno (14) observed that while the NDSs had guidance on involving people living with dementia (PLwD) and their caregivers’ involvement in future plans, the strategies differed in their approach to involving PLwD in advanced stages and care for minority and disadvantaged groups, and people with early onset dementia.

The analysis of the text of NDSs selected for this study also shows that the major emphasis is on care, which is confirmed by other studies too. Over and above focusing on “care”, there have been suggestions for NDSs to focus on increasing awareness and reducing the stigma surrounding dementia, as well as promoting research on dementia (7,8,14). It may be noted that these studies are undertaken in the context of countries around the world, and they do not specifically study European countries. Hampel et al. (47) observed that the dementia strategies focus on risk reduction / prevention and early diagnosis. This implies that the NDSs of European countries are required to focus more on provisions to deal with stigma and increase dementia awareness, as well as about early diagnosis and prevention, to match their importance with care for PLwD.

Alzheimer Europe (15) findings suggest that the most NDSs focus on the provision of health and social care services for people with dementia, diagnosis and treatment, and the training of practitioners. National dementia strategies also focus on raising awareness about dementia and undertaking research on dementia. Dementia strategies of some countries also focused on dementia prevention. It may be noted that in this study, the key focus areas are identified based on frequency of occurrence of a term or a combination of terms in the NDSs clusters. This in no way implies that there is no provision for other areas like prevention, raising awareness etc. It only means that other areas require higher attention that what is already accorded in the NDSs. On the other hand, the findings of Alzheimer Europe (16) show that while countries have been proactive in publishing the policies, little efforts have been made towards allocating finance for implementing these policies.

Countries also need to devise a set of dementia indicators, on which, data can be collected every every two years, as suggested in the GAD.

A lot has already been done for PLwD and their caregivers. Yet, a lot is required to be done for prevention and PCC. We recommend prevention and PCC to be accorded the highest priority, after care, in the NDSs. Awareness and destigmatising dementia, which are necessary for their right to live with dignity should also be accorded high priority.

## 5. Conclusion

We began with the reference to the Glasgow Declaration and the Global Action Plan of the WHO, and their key focus areas, and then compared the clusters formed of national dementia strategies of 15 European countries. The results suggest that the dementia strategies are highly biased towards care provision for the PLwD. Care is undoubtedly, one of the crucial aspects, given the prevalence and growth in the number of persons with dementia. However, as suggested in the GAP, dementia prevention, person-centered care, increasing awareness, and reducing stigma around dementia, plus, devising a set of dementia indicators and regular collection of data on those indicators require attention.

## Data Availability

The data underlying the results presented in the study are available from the website of Alzheimer Europe: https://www.alzheimer-europe.org/policy/national-dementia-strategies

## Acknowledgements

Smruti Bulsari, University of Essex, is supported by the National Institute for Health and Care Research – Applied Research Collaboration East of England (NIHR ARC EoE) and the Alzheimer’s Society, funded through a Post-Doctoral Fellowship. The NIHR ARC EoE is hosted by Cambridgeshire and Peterborough NHS Foundation Trust. The views expressed are those of the authors and not necessarily those of the NIHR or the Department of Health and Social Care.

## Notes

### Competing Interest Statement

The authors have declared no competing interest.

### Funding Statement

Yes

### Author Declarations

None. The research analyses only the published documents, available in public domain.

## References

1. WHO. Dementia: Key Facts. 2023 [cited 2024 Apr 15]; Available from: https://www.who.int/news-room/fact-sheets/detail/dementia

2. Ali GC, Guerchet M, Wu YT, Prina M. The Global Prevalence of Dementia. In: World Alzheimer Report 2015: The Global Impact of Dementia: An analysis of prevalence, incidence, cost and trends [Internet]. London: Alzheimer’s Disease International and Bupa; 2015. Available from: https://www.alzint.org/u/WorldAlzheimerReport2015.pdf

3. Long S, Benoist C, Weidner W. World Alzheimer Report 2023: : Reducing dementia risk: never too early, never too late [Internet]. London: Alzheimer’s Disease International; 2023. Available from: https://www.alzint.org/u/World-Alzheimer-Report-2023.pdf

4. Prince M, Jackson J. World Alzheimer Report 2009 [Internet]. London: Alzheimer’s Disease International; 2009. Available from: https://www.alzint.org/u/WorldAlzheimerReport.pdf

5. WHO. Global Action Plan on the Public Health Response to Dementia [Internet]. Geneva; 2017. Available from: https://iris.who.int/bitstream/handle/10665/259615/9789241513487-eng.pdf?sequence=1

6. Alzheimer Europe. National Dementia Strategies [Internet]. Policy. 2024 [cited 2024 Feb 29]. Available from: https://www.alzheimer-europe.org/policy/national-dementia-strategies

7. Edick C, Holland N, Ashbourne J, Elliott J, Stolee P. A review of Canadian and international dementia strategies. Healthc Manage Forum. 2017 Jan 1;30(1):32–9.

8. Chow S, Chow R, Wan A, Lam HR, Taylor K, Bonin K, et al. National Dementia Strategies: What Should Canada Learn? Can Geriatr J CGJ. 2018 Jun;21(2):173–209.

9. Alzheimer’s Disease International. Dementia Plans [Internet]. Policy. 2024 [cited 2024 Apr 12]. Available from: https://www.alzint.org/what-we-do/policy/dementia-plans/

10. Nakanishi M, Nakashima T, Shindo Y, Miyamoto Y, Gove D, Radbruch L, et al. An evaluation of palliative care contents in national dementia strategies in reference to the European Association for Palliative Care white paper. Int Psychogeriatr. 2015/02/13 ed. 2015;27(9):1551–61.

11. Fortinsky RH, Downs M. Optimizing Person-Centered Transitions In The Dementia Journey: A Comparison Of National Dementia Strategies. Health Aff (Millwood). 2014 Apr 1;33(4):566–73.

12. Schmachtenberg T, Monsees J, Hoffmann W, van den Berg N, Stentzel U, Thyrian JR. Comparing national dementia plans and strategies in Europe – is there a focus of care for people with dementia from a migration background? BMC Public Health. 2020 May 26;20(1):784.

13. Nakanishi M, Nakashima T. Features of the Japanese national dementia strategy in comparison with international dementia policies: How should a national dementia policy interact with the public health- and social-care systems? Alzheimers Dement. 2014 Jul 1;10(4):468–476.e3.

14. Vinay R, Biller-Andorno N. A critical analysis of national dementia care guidances. Health Policy. 2023 Apr 1;130:104736.

15. Alzheimer Europe. Dementia in Europe Yearbook 2018: Comparison of National Dementia Strategies in Europe [Internet]. Luxembourg; 2018 [cited 2024 Apr 12]. Available from: https://www.alzheimer-europe.org/sites/default/files/alzheimer_europe_dementia_in_europe_yearbook_2018.pdf

16. Alzheimer Europe. European Dementia Monitor 2020: Comparing and Benchmarking National Dementia Strategies and Policies. Luxembourg; 2020.

17. Alzheimer Europe. Glasgow Declaration [Internet]. 2014 [cited 2023 Oct 31]. Available from: https://www.alzheimer-europe.org/policy/campaign/glasgow-declaration-2014#:~:text=The%20Glasgow%20Declaration%20called%20for,global%20action%20plan%20on%20dementia.

18. Cahill S. WHO’s global action plan on the public health response to dementia: some challenges and opportunities. Aging Ment Health. 2020 Feb 1;24(2):197–9.

19. Arthurton L, Lynch C, Weidner WS. From Plan to Impact-progress towards targets of the WHO Global action plan on dementia. Alzheimers Dement. 2022;18:e061449.

20. Hofferbert RI, Cingranelli DL. 593Public Policy and Administration: Comparative Policy Analysis. In: Goodin RE, Klingemann HD, editors. A New Handbook of Political Science [Internet]. Oxford University Press; 1998 [cited 2024 Dec 4]. p. 0. Available from: 10.1093/0198294719.003.0025

21. Dogaru Cruceanu TC. The comparative method for policy studies: the thorny aspects. HOLISTICA – J Bus Public Adm. 2019;10(1):56–67.

22. Peters BG. The comparative method and comparative policy analysis. In: Handbook of Research Methods and Applications in Comparative Policy Analysis [Internet]. Cheltenham, UK: Edward Elgar Publishing; 2020. p. 20–32. Available from: https://www.elgaronline.com/view/edcoll/9781788111188/9781788111188.00007.xml

23. Hagen L, Harrison TM, Uzuner Ö, Fake T, Lamanna D, Kotfila C. Introducing Textual Analysis Tools for Policy Informatics: A Case Study of e-Petitions. In: Proceedings of the 16th Annual International Conference on Digital Government Research [Internet]. New York, NY, USA: Association for Computing Machinery; 2015. p. 10–9. (dg.o ’15). Available from: 10.1145/2757401.2757421

24. Safaei M, Longo J. The End of the Policy Analyst? Testing the Capability of Artificial Intelligence to Generate Plausible, Persuasive, and Useful Policy Analysis. Digit Gov Res Pr [Internet]. 2023 Aug; Available from: 10.1145/3604570

25. Smith TB, Vacca R, Mantegazza L, Capua I. Natural language processing and network analysis provide novel insights on policy and scientific discourse around Sustainable Development Goals. Sci Rep. 2021 Nov 17;11(1):22427.

26. Liu Bi, Li X, Lee WS, Yu PS. Text Classification by Labeling Words. Assoc Adv Artif Intell AAAI. 2004;4:425–30.

27. Heimerl F, Lohmann S, Lange S, Ertl T. Word Cloud Explorer: Text Analytics Based on Word Clouds. In: 2014 47th Hawaii International Conference on System Sciences. 2014. p. 1833–42.

28. Jurafsky D, Martin JH. N-gram Language Models. In: Speech and Language Processing: An Introduction to Natural Language Processing, Computational Linguistics, and Speech Recognition [Internet]. 3rd ed. Online; 2018 [cited 2024 Apr 15]. Available from: https://github.com/yanshengjia/ml-road/blob/master/resources/Speech%20and%20Language%20Processing%20(3rd%20Edition).pdf

29. Wallace LS. A view of health care around the world. Ann Fam Med. 2013 Feb;11(1):84.

30. Bholowalia P, Kumar A. EBK-Means: A Clustering Technique based on Elbow Method and K-Means in WSN. Int J Comput Appl. 2014;105(9):17–24.

31. Umargono E, Suseno JE, Vincensius Gunawan SK. K-Means Clustering Optimization using the Elbow Method and Early Centroid Determination Based-on Mean and Median. In: CONRIST. Yogyakarta, Indonesia: SciTePress; 2019. p. 234–40.

32. Theodorou M, Charalambous C, Petrou C, Cylus J. Health system Review. Cyprus Health Syst Transit. 2012;14(6):1–128.

33. HM of Government of Gibraltar. Health [Internet]. 2024 [cited 2024 Apr 19]. Available from: https://www.gibraltar.gov.gi/health

34. Rosen B, Waitzberg R, Merkur S. Israel: Health System Review. Health Syst Transit. 2015;17(6):1–212.

35. Cantarero-Prieto D, Leon PL, Blazquez-Fernandez C, Juan PS, Cobo CS. The economic cost of dementia: A systematic review. Dementia. 2020 Nov 1;19(8):2637–57.

36. Connolly S, Gillespie P, O’Shea E, Cahill S, Pierce M. Estimating the economic and social costs of dementia in Ireland. Dementia. 2014 Jan 1;13(1):5–22.

37. Deb A, Thornton JD, Sambamoorthi U, Innes K. Direct and indirect cost of managing alzheimer’s disease and related dementias in the United States. Expert Rev Pharmacoecon Outcomes Res. 2017 Apr;17(2):189–202.

38. Livingston G, Huntley J, Sommerlad A, Ames D, Ballard C, Banerjee S, et al. Dementia prevention, intervention, and care: 2020 report of the Lancet Commission. The Lancet. 2020 Aug 8;396(10248):413–46.

39. Livingston G, Huntley J, Liu KY, Costafreda SG, Selbæk G, Alladi S, et al. Dementia prevention, intervention, and care: 2024 report of the Lancet standing Commission. The Lancet. 2024 Aug 10;404(10452):572–628.

40. Braun A, Höfler M, Auer S. Cost-Effectiveness of Prevention for People at Risk for Dementia: A Scoping Review and Qualitative Synthesis. J Prev Alzheimers Dis. 2024 Mar 1;11(2):402–13.

41. Wimo A, Handels R, Antikainen R, Eriksdotter M, Jönsson L, Knapp M, et al. Dementia prevention: The potential long-term cost-effectiveness of the FINGER prevention program. Alzheimers Dement. 2023 Mar 1;19(3):999–1008.

42. McRae I, Zheng L, Bourke S, Cherbuin N, Anstey KJ. Cost-Effectiveness of Dementia Prevention Interventions. J Prev Alzheimers Dis. 2021 Apr 1;8(2):210–7.

43. Tay FHE, Thompson CL, Nieh CM, Nieh CC, Koh HM, Tan JJC, et al. Person-centered care for older people with dementia in the acute hospital. Alzheimers Dement Transl Res Clin Interv. 2018 Jan 1;4:19–27.

44. Chenoweth L, Williams A, Fry M, Endean E, Liu Z. Outcomes of Person-centered Care for Persons with Dementia in the Acute Care Setting: A Pilot Study. Clin Gerontol. 2022 Aug 8;45(4):983–97.

45. Evans GW. Output and Unemployment Dynamics in the United States: 1950-1985. J Appl Econom. 1989;4(3):213–37.

46. Mohr W, Rädke A, Afi A, Edvardsson D, Mühlichen F, Platen M, et al. Key Intervention Categories to Provide Person-Centered Dementia Care: A Systematic Review of Person-Centered Interventions. J Alzheimers Dis. 2021;84(1):343–66.

47. Hampel H, Vergallo A, Iwatsubo T, Cho M, Kurokawa K, Wang H, et al. Evaluation of major national dementia policies and health-care system preparedness for early medical action and implementation. Alzheimers Dement. 2022 Oct 1;18(10):1993–2002.

